# Randomized controlled trial transfusing convalescent plasma as post-exposure prophylaxis against SARS-CoV-2 infection

**DOI:** 10.1101/2021.12.13.21267611

**Authors:** Shmuel Shoham, Evan M Bloch, Arturo Casadevall, Daniel Hanley, Bryan Lau, Kelly Gebo, Edward Cachay, Seble G. Kassaye, James H. Paxton, Jonathan Gerber, Adam C Levine, Judith Currier, Bela Patel, Elizabeth S. Allen, Shweta Anjan, Lawrence Appel, Sheriza Baksh, Paul W. Blair, Anthony Bowen, Patrick Broderick, Christopher A Caputo, Valerie Cluzet, Marie Elena Cordisco, Daniel Cruser, Stephan Ehrhardt, Donald Forthal, Yuriko Fukuta, Amy L. Gawad, Thomas Gniadek, Jean Hammel, Moises A. Huaman, Douglas A. Jabs, Anne Jedlicka, Nicky Karlen, Sabra Klein, Oliver Laeyendecker, Karen Lane, Nichol McBee, Barry Meisenberg, Christian Merlo, Giselle Mosnaim, Han-Sol Park, Andrew Pekosz, Joann Petrini, William Rausch, David M. Shade, Janna R. Shapiro, J. Robinson Singleton, Catherine Sutcliffe, David L. Thomas, Anusha Yarava, Martin Zand, Jonathan M. Zenilman, Aaron A.R. Tobian, David Sullivan

## Abstract

**BACKGROUND:** The efficacy of SARS-CoV-2 convalescent plasma (CCP) for preventing infection in exposed, uninfected individuals is unknown. We hypothesized that CCP might prevent infection when administered before symptoms or laboratory evidence of infection.

**METHODS:** This double-blinded, phase 2 randomized, controlled trial (RCT) compared the efficacy and safety of prophylactic high titer (≥1:320) CCP with standard plasma. Asymptomatic participants aged ≥18 years with close contact exposure to a person with confirmed COVID-19 in the previous 120 hours and negative SARS-CoV-2 test within 24 hours before transfusion were eligible. The primary outcome was development of SARS-CoV-2 infection.

**RESULTS:** 180 participants were enrolled; 87 were assigned to CCP and 93 to control plasma, and 170 transfused at 19 sites across the United States from June 2020 to March 2021. Two were excluded for SARS-CoV-2 RT-PCR positivity at screening. Of the remaining 168 participants, 12/81 (14.8%) CCP and 13/87 (14.9%) control recipients developed SARS-CoV-2 infection; 6 (7.4%) CCP and 7 (8%) control recipients developed COVID-19 (infection with symptoms). There were no COVID-19-related hospitalizations in CCP and 2 in control recipients. There were 28 adverse events in CCP and 58 in control recipients. Efficacy by restricted mean infection free time (RMIFT) by 28 days for all SARS-CoV-2 infections (25.3 vs. 25.2 days; p=0.49) and COVID-19 (26.3 vs. 25.9 days; p=0.35) were similar for both groups.

**CONCLUSION:** In this trial, which enrolled persons with recent exposure to a person with confirmed COVID-19, high titer CCP as post-exposure prophylaxis appeared safe, but did not prevent SARS-CoV-2 infection.

**Trial Registration:** Clinicaltrial.gov number NCT04323800.

## Introduction

Severe acute respiratory syndrome coronavirus 2 (SARS-CoV-2) is responsible for Coronavirus Disease 2019 (COVID-19) and the pandemic that has claimed millions of lives^1^. Especially at the pandemic’s onset, effective preventive strategies were limited. Even by late 2021, only half of the world’s population has been vaccinated, and some do not respond to vaccination. ^2 3^. The urgency of effective prevention is highest within households of SARS-CoV-2 infected persons since 10-50% will be secondarily infected. Passive immunotherapy using preformed antibodies is effective as post-exposure prophylaxis (PEP) against many infections^4–7^. Combinations of monoclonal antibodies (mAb) are effective as COVID-19 PEP ^8,9^. COVID-19 convalescent plasma (CCP) also may confer protection during early infection and in those without antibodies ^11–13^. CCP has some advantages over mAb’s, including ease of procurement, low cost, and resilience against viral variants^14^. This study sought to evaluate the safety and efficacy of CCP containing high titers of anti-SARS-CoV-2 antibodies as PEP.

## Methods

### Study design and overview

A randomized, double-blind, placebo-controlled clinical trial was conducted from to compare the safety and efficacy of transfusion of CCP (intervention) with SARS-CoV-2 non-immune control plasma.

### Participants

Asymptomatic participants aged ≥18 years who had a close contact exposure to a person with confirmed COVID-19 in the previous 120 hours and did not have SARS-CoV-2 vaccination, and past or active SARS-CoV-2 infection were eligible. Transfused participants positive by RT-PCR at screening were excluded from analyses. Participants were enrolled at 19 United States centers between June 11, 2020 to June 23, 2021

### Randomization to treatment arm and masking

Eligible subjects were randomized 1:1 to receive 1 unit of CCP or 1 unit of control plasma using interactive web-based systems. CCP and control plasma were in standard plasma bags, with identical labels.

### Intervention

CCP donors were eligible for collection if they had a history of a positive molecular assay test result for SARS-CoV-2 infection, met standard criteria for blood donation, and had SARS-CoV-2 antibody levels ≥ 1:320 titer by Euroimmun ELISA, [Mountain Lakes, NJ] at screening. Subsequent to an FDA Emergency Use Authorization (February, 4, 2021), CCP was only used if it met the 1:320 dilutional titer criterion and an Arbitrary Unit of 3.5 at a 1:101 dilution by Euroimmun IgG ELISA. Control was standard SARS-CoV-2 non-immune plasma collected before January 1, 2020, or seronegative for SARS-CoV-2.

### Primary Outcome

The primary efficacy outcome was incident SARS-CoV-2 infection by study day 28 by positive RT-PCR testing conducted on collected nasal swabs or by clinical RT-PCR testing conducted outside the study

Individuals were followed for 90 days with visits at days 0 (transfusion), 1, 3, 7, 14, 28, 60, and 90. Nasal swabs were collected at screening (days −1 to 0) and at days 1, 7, 14, and 28. Assessments for COVID-19 (symptomatic infection) were conducted at screening, transfusion (day 0), and days 1, 3, 7, 14, 28, and 60. Viral testing was performed using RT-PCR that targeted the SARS-CoV-2 nucleocapsid gene.

### Secondary efficacy outcomes

Disease severity was measured to day 28 using a clinical event scale and evaluated using an ordinal logistic model. Efficacy for preventing SARS-CoV-2 infection and COVID-19 was examined based on donor antibody titer through characterization of donor IgG, including end point titers and area under the curve (AUC) using a standardized ELISA to measure IgG against the spike and receptor binding proteins and anti-SARS-CoV-2 IgG against recombinant S1 domain of the SARS-CoV-2 spike protein (Euroimmun) as previously described^15^.

### Safety assessments

Reportable adverse events (AEs) included serious AEs (SAEs) and transfusion reactions. An independent safety monitor, masked to randomized assignment, reviewed all AEs, SAEs and changes in baseline safety laboratory values.

### Data management and statistical analyses

The pre-specified primary analysis of cumulative SARS-CoV-2 infection was conducted using a time-to-event analysis to compare the restricted mean survival time, referred to henceforth as restricted mean infection free time (RMIFT).

We calculated and compared the restricted mean survival times by 28 days and risk difference (RD) by treatment arm in a modified intention to treat (mITT) analysis. We performed the primary analysis according to the participants’ original randomized treatment groups excluding those who did not receive a transfusion of study plasma and those who were later found to have been test positive at transfusion ^16^. Analyses were adjusted for variables potentially related to the outcome in order to increase estimate precision^16^. Demographic and clinical variables were measured at baseline. To determine which pre-specified candidate variables to include, we conducted variable selection by random survival forest in the entire sample (i.e., not including an indicator term for treatment arm) and masked to treatment allocation. This algorithm was implemented on the mITT sample to identify the prognostic baseline variables for the entire sample.

Baseline characteristics are reported as proportions or medians with interquartile ranges (IQR) for continuous variables. Time-to-event analysis was computed from the time of transfusion until development of a positive molecular test for infection. Analyses were repeated using only clinical illness with COVID-19 as the outcome. Targeted minimum loss-based estimation (TMLE) was used for difference in RMIFT by 28 days and risk of infection. Time scale was days from transfusion. A one-sided test with type I error of 0.05 was used to determine statistical significance.

A secondary outcome was disease severity by day 28 using a clinical event scale ranging from no infection to death. The most severe status by the day 28 visit was ascertained using a TMLE estimator for ordinal outcomes and adjusted for the pre-specified candidate variables selected by the algorithmic approach.^17,18^

A pre-specified sensitivity analysis was restricted to participants who remained infection-free up to day 4 to allow for potential lag between transfusion and effect from passive antibody transfer.

### Donor antibody titers

Analysis for donor antibody titer was conducted for AUC as a continuous variable: controls were assigned a value of zero. To model antibody effect, a flexible Weibull time to event model was used^19^ to estimate the hazard ratios. To allow for non-linearity, both natural cubic splines and fractional polynomials were assessed choosing the model with the lowest Akaike Information Criterion (AIC)^20^.

### Safety

Rates of severe transfusion reactions, AEs, grade 3 or 4 AEs, and death were evaluated by treatment arm; 95% confidence intervals (CI) were calculated using skewness-corrected asymptotic score for exact CI^21^, using the R package ‘ratesci’.

### Conditional Power Analysis

The trial did not meet the target sample of 500 participants as enrollment stopped with widespread vaccine availability. The sample size calculation is provided as supplementary material. A conditional power analysis, using the R package ‘gsDesign’, was conducted to assess the likelihood of providing evidence for the efficacy of convalescent plasma.

### Ethical Review and Trial Oversight

Approval was obtained from the Institutional Review Boards at Johns Hopkins University School of Medicine functioning as single IRB for all participating sites. An independent data and safety monitoring board provided oversight and reviewed efficacy and safety as the study was conducted. All participants provided written informed consent.

## Results

Of 1,138 participants screened, 180 (15.8%) were eligible and consented to the study and 170 were transfused (82 CCP; 88 control plasma; Figure 1). Of those transfused, two were excluded from efficacy analyses for baseline SARS-CoV-2 RT-PCR positivity. Table 1 lists participants’ demographic and baseline characteristics. Median time from exposure to transfusion was 2 days (IQR 1-4). Seven participants (3 CCP recipients) did not complete all study components.

**Table 1:**
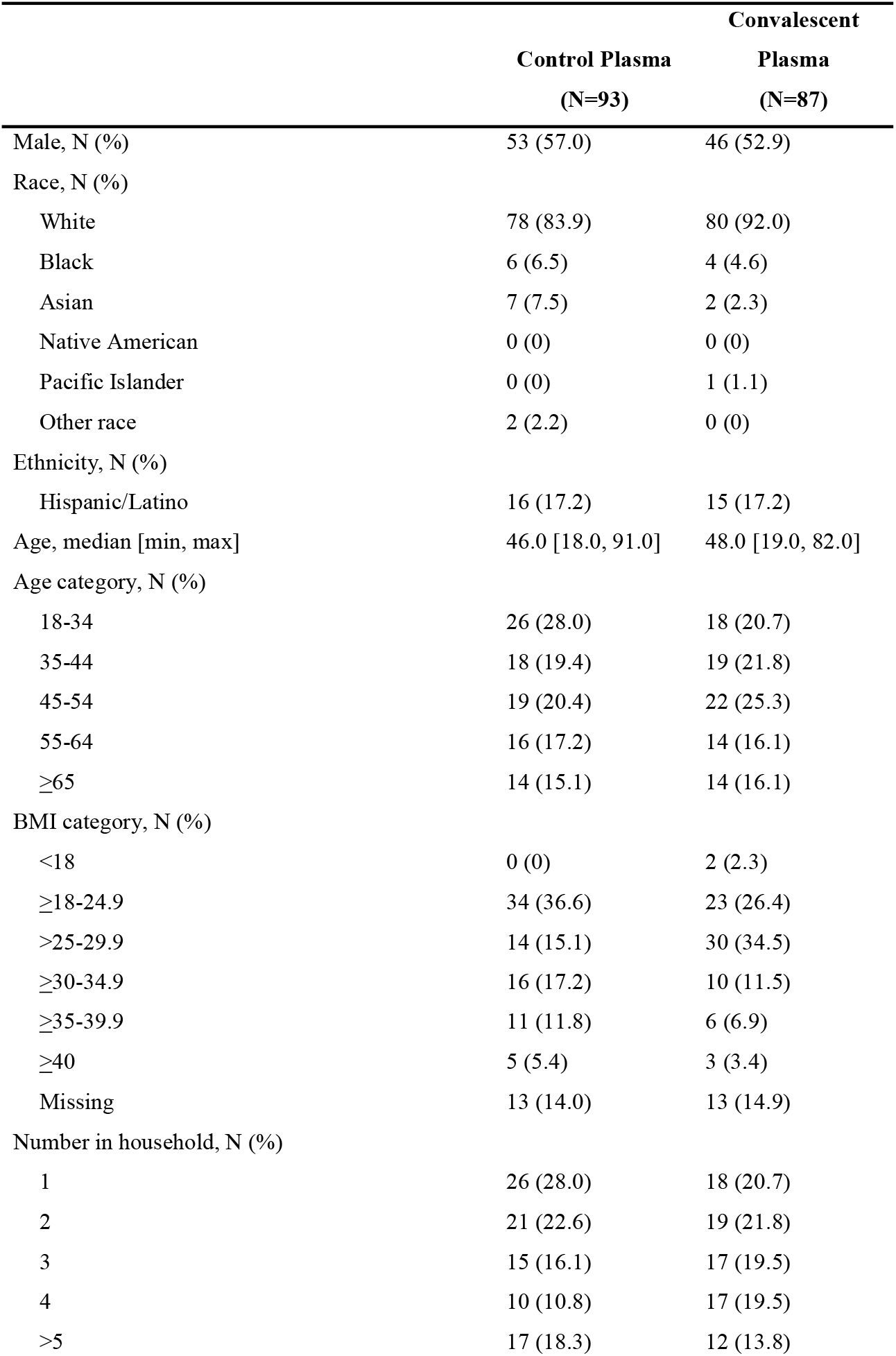
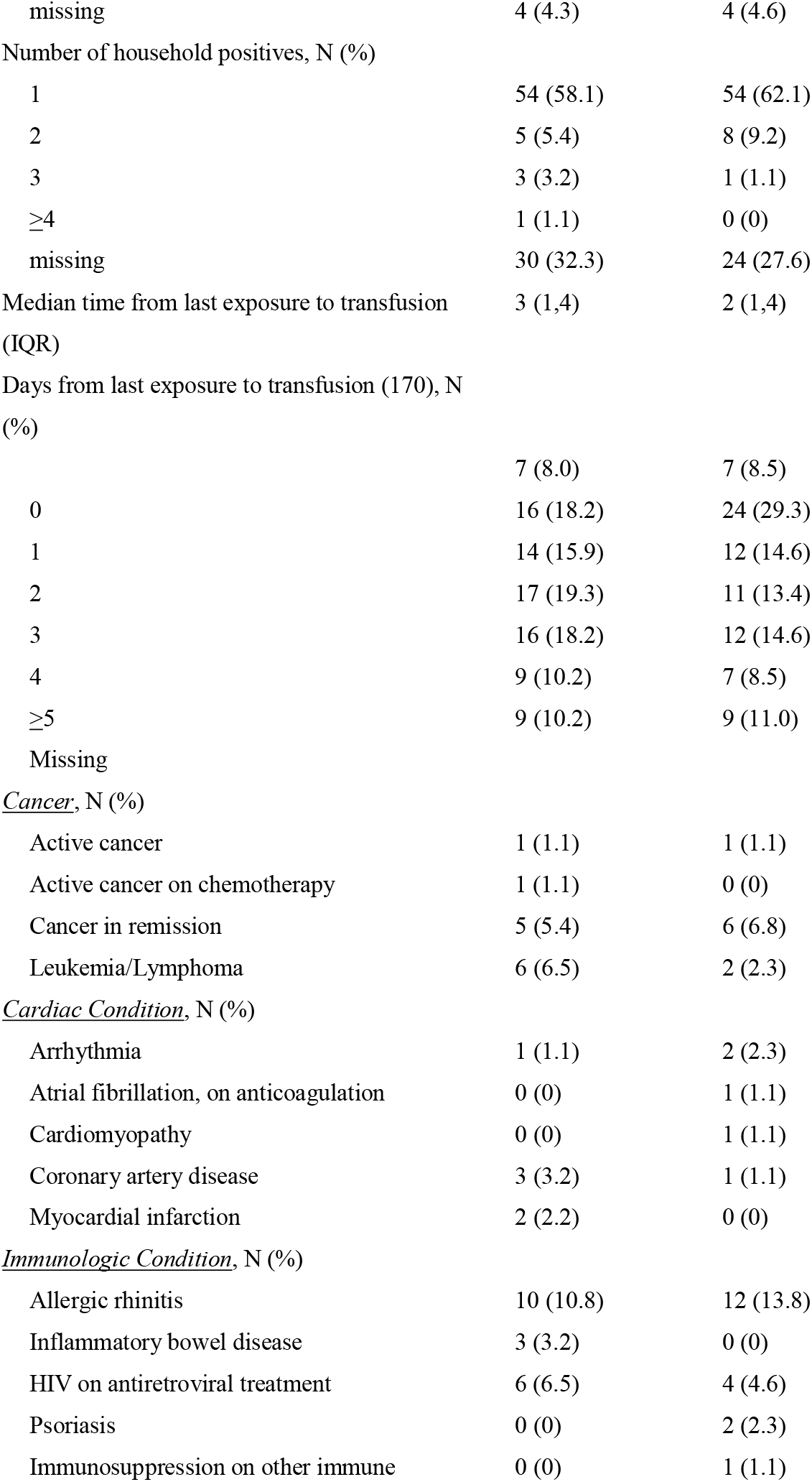
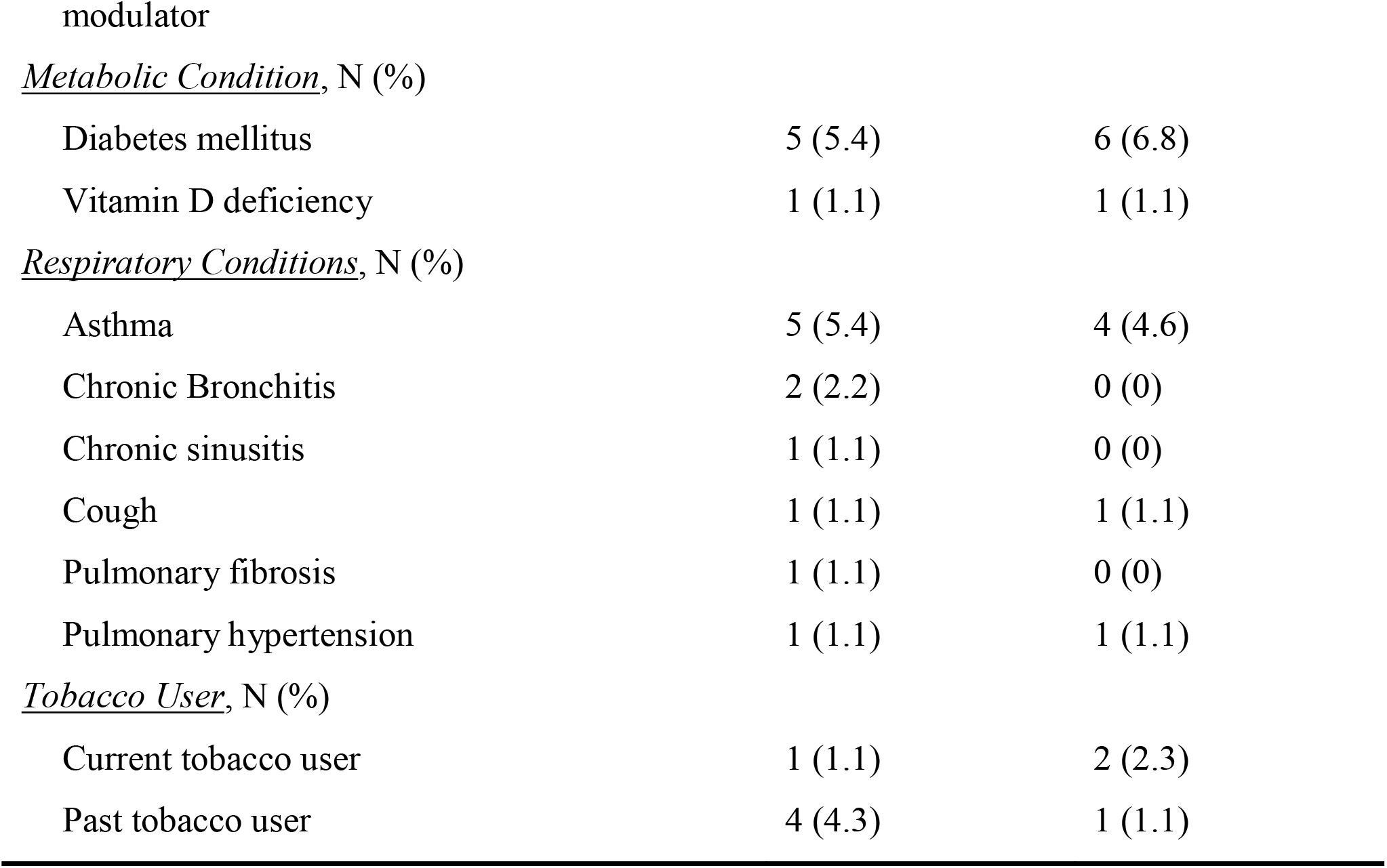
Demographics and Medical Conditions at Randomization.

**Figure 1:**
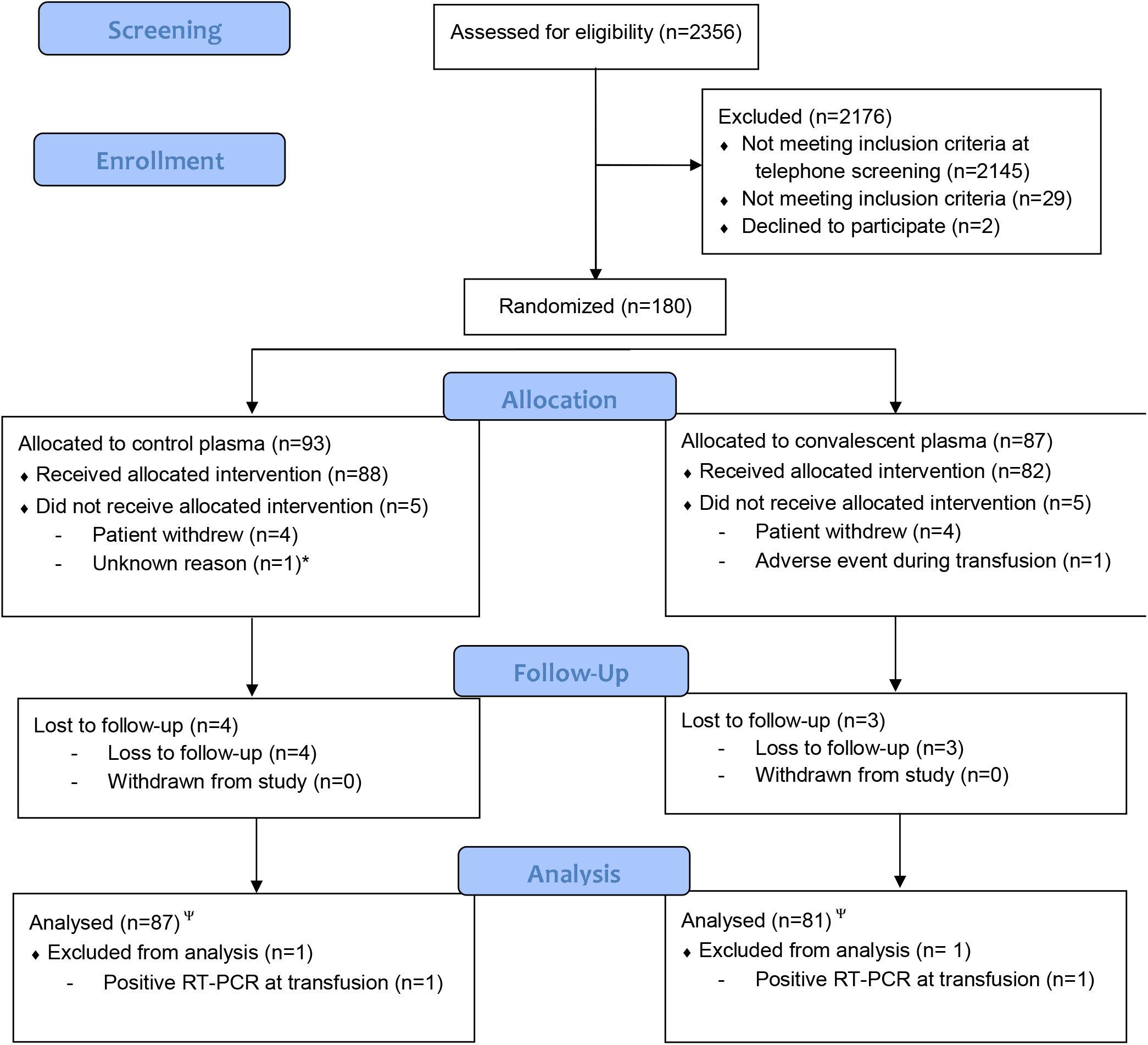
Consort Diagram: ^Ψ^ Intention to treat analysis, including all transfused individuals. Those lost to follow-up between transfusion to end of follow-up contributed to the time at risk. Individuals with positive RT-PCR on day of transfusion were removed from analysis. * One randomized participant was found ineligible after randomization.

CCP from 70 unique donations was transfused to 82 recipients; the anti-S IgG inverse endpoint titers were > 1,000 except for a single unit at 540. 87% were hospital qualified EUA high titer by Euroimmun ratio ≥3.5. Virus neutralizing antibody titers ranged from 1:20 to 1:640 and were present in 94% of 50 tested donors.

### Primary Outcomes

Of the 168 participants in the mITT analyses, 12/81 (14.8%) CCP and 13/87 (14.9%) control recipients tested positive for SARS-CoV-2 RNA. Three were positive on day 1 post-transfusion and 3 on days 2-3. Cumulative incidence of confirmed infections and differences in RMIFT are shown in figures 2a,c,e. Analyses excluding the six infections occurring through day 3 are shown in figures 2b,d,f. The RMIFT by 28 days was 25.3 days for CCP and 25.2 for control recipients (p=0.47. The RD was 0.01 (p=0.42) lower for CCP. Excluding infections through day 3 the RMIFT was 26.6 days for CCP and 25.8 for control recipients (p=0.15). The RD was 0.04 (p=0.21) lower for CCP.

**Figure 2:**
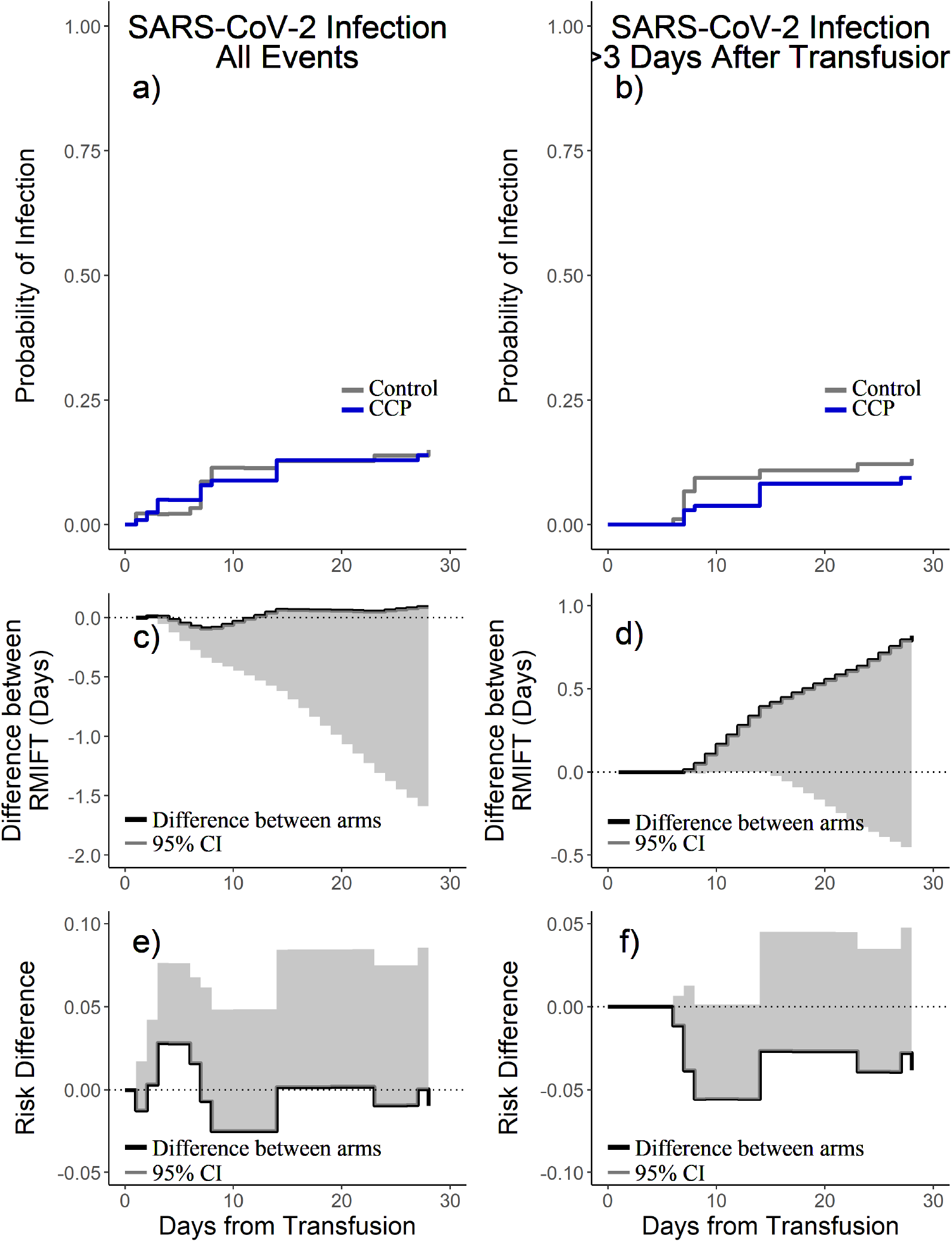
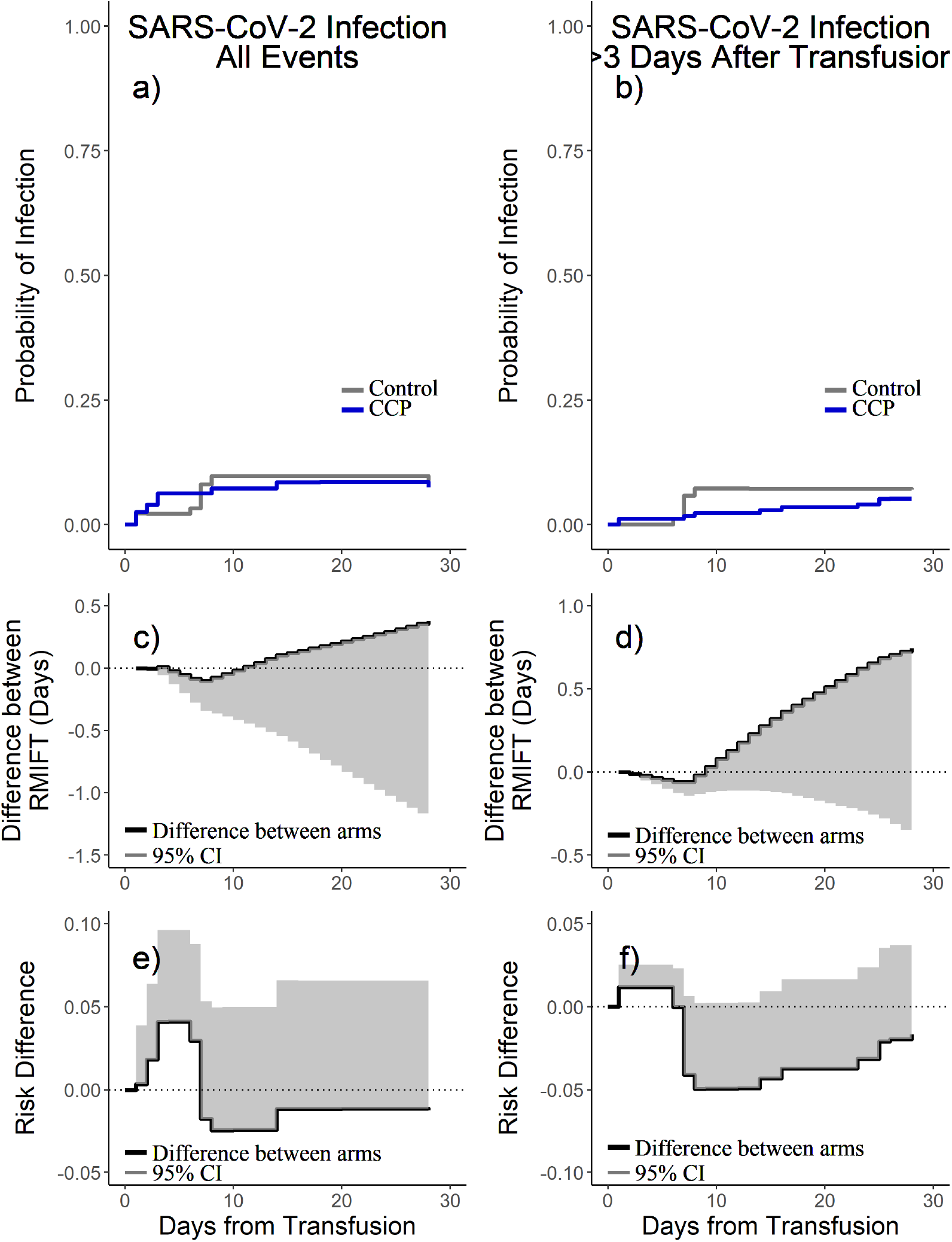
a, b: Cumulative incidence of laboratory detected SARS-CoV-2 infection; c.d: difference in restricted mean infection free time (RMIFT) (> 0: increased expected days to infection for CCP); e, f: risk difference (lower panels, < 0: lower risk of infection for CCP). 95% CI=One-sided 95% confidence interval.

Six (7.4%) CCP and 7 (8%) control recipients had COVID-19 (4 and 5 after day 3 from transfusion). Cumulative incidence of COVID-19 and differences in RMIFT are shown in figure 3a,c,e. Analyses excluding infections through day 3 are shown in figures 3b,d,f. The RMIFT by 28 days was 26.3 for the CCP and 25.9 days for the control recipients. The RD between groups was 0.012 lower for CCP. Excluding infections through day 3, the CCP group was consistently, but not significantly, better than control (difference in RMIFT =0.7 days, p=0.14; RD=0.017).

**Figure 3:**
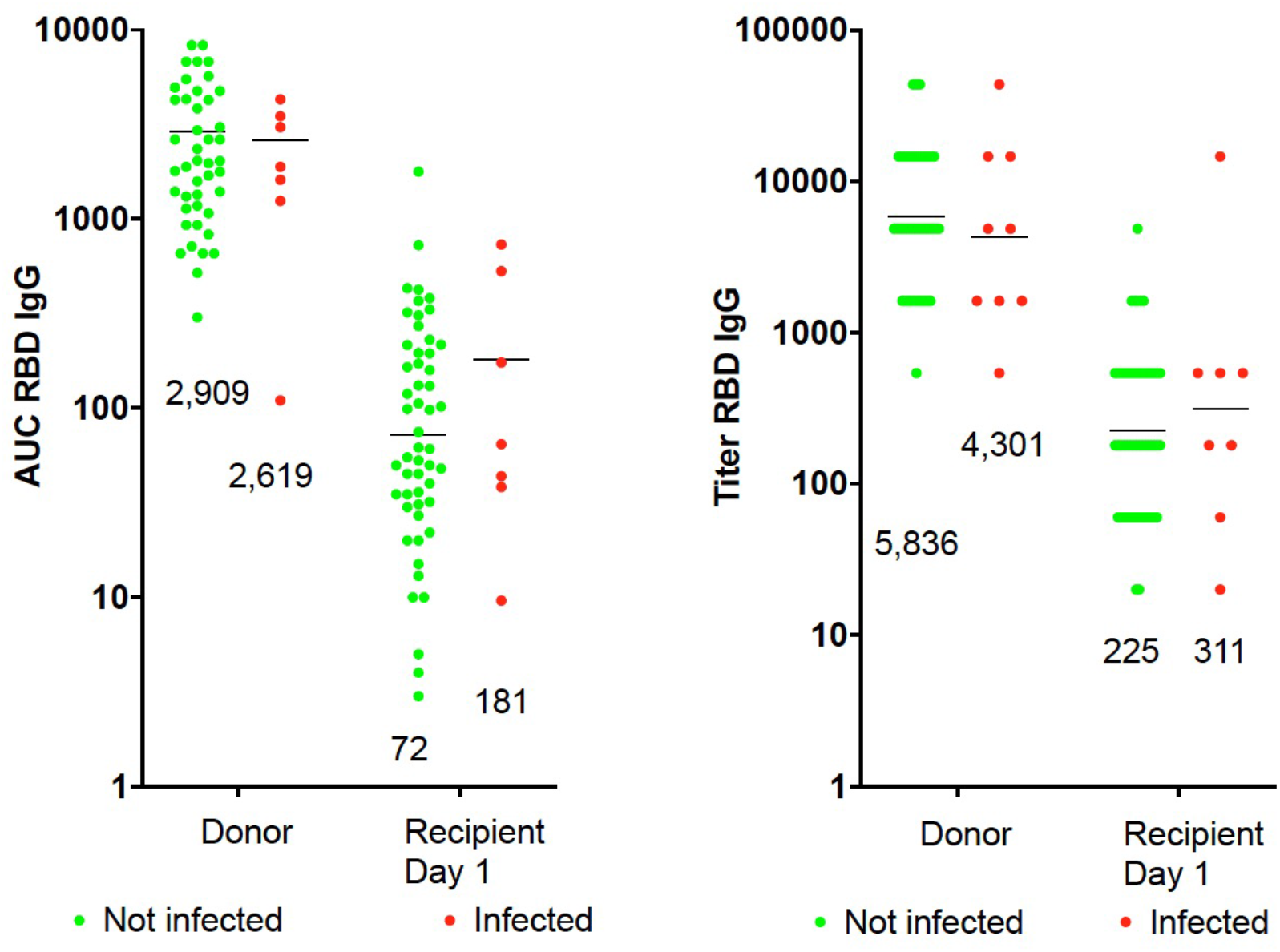
a, b: Cumulative incidence of COVID-19; c, d: difference in restricted mean infection free time (RMIFT) (> 0: increased expected days to infection for CCP); e, f: risk difference (< 0: lower risk of infection for CCP). 95% CI=One-sided 95% confidence interval.

Conditional power analyses were conducted since the target enrollment (500 transfused) was not reached. Had target enrollment been reached it is unlikely that statistically significant results would have been achieved, with chances for significant differences in RMIFT and RD calculated as 0.3% and 0.6% respectively.

### Adverse Events

There were 86 reported AEs, of which 58 occurred with CCP and 28 with control plasma; 17/86 events were grade 3 or 4 Five participants required hospitalization (2 for COVID-19) all with control plasma (Supplemental Materials). CCP recipients had a lower proportion of any AEs (p=0.005), and severe AEs (p=0.06) (Table 2).

**Table 2:**
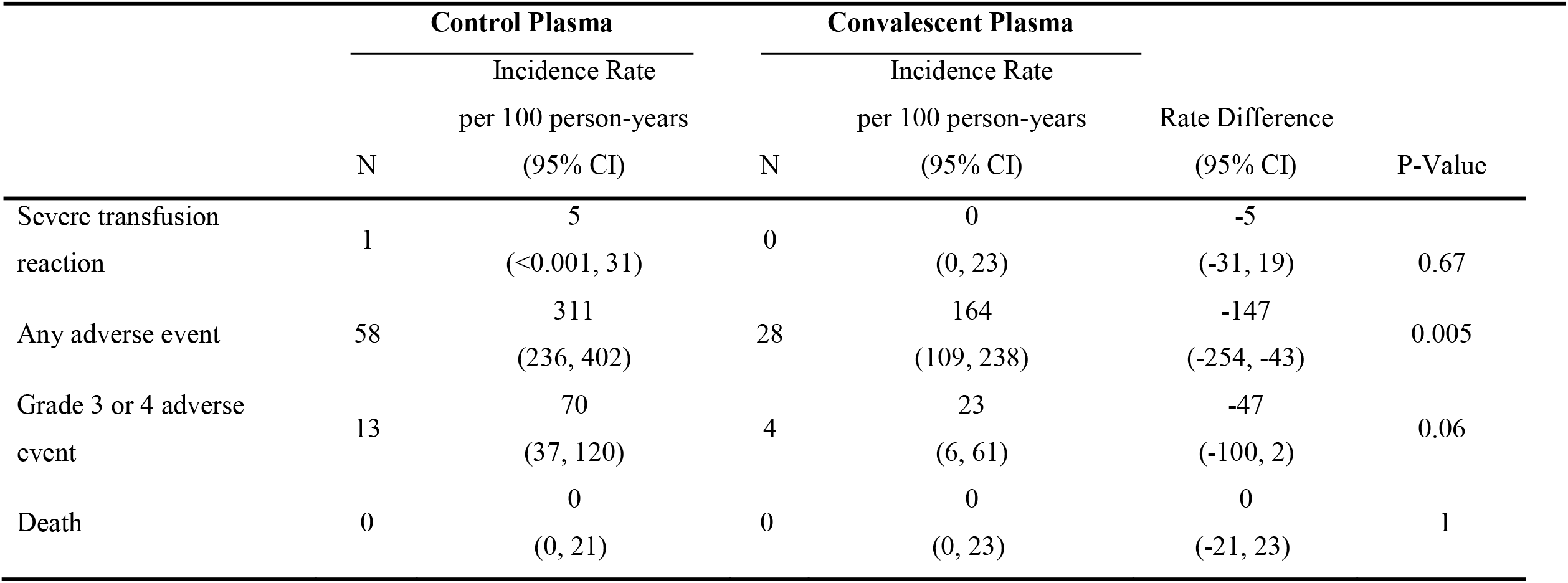
Adverse events.

### Clinical Severity Score

Two participants required hospitalizations due to COVID-19. Both were control recipients (Table 3). The distribution of clinical severity was similar between the two groups for all events after transfusion (OR 0.99) and for events >3 days after transfusion (OR 0.94).

**Table 3:**
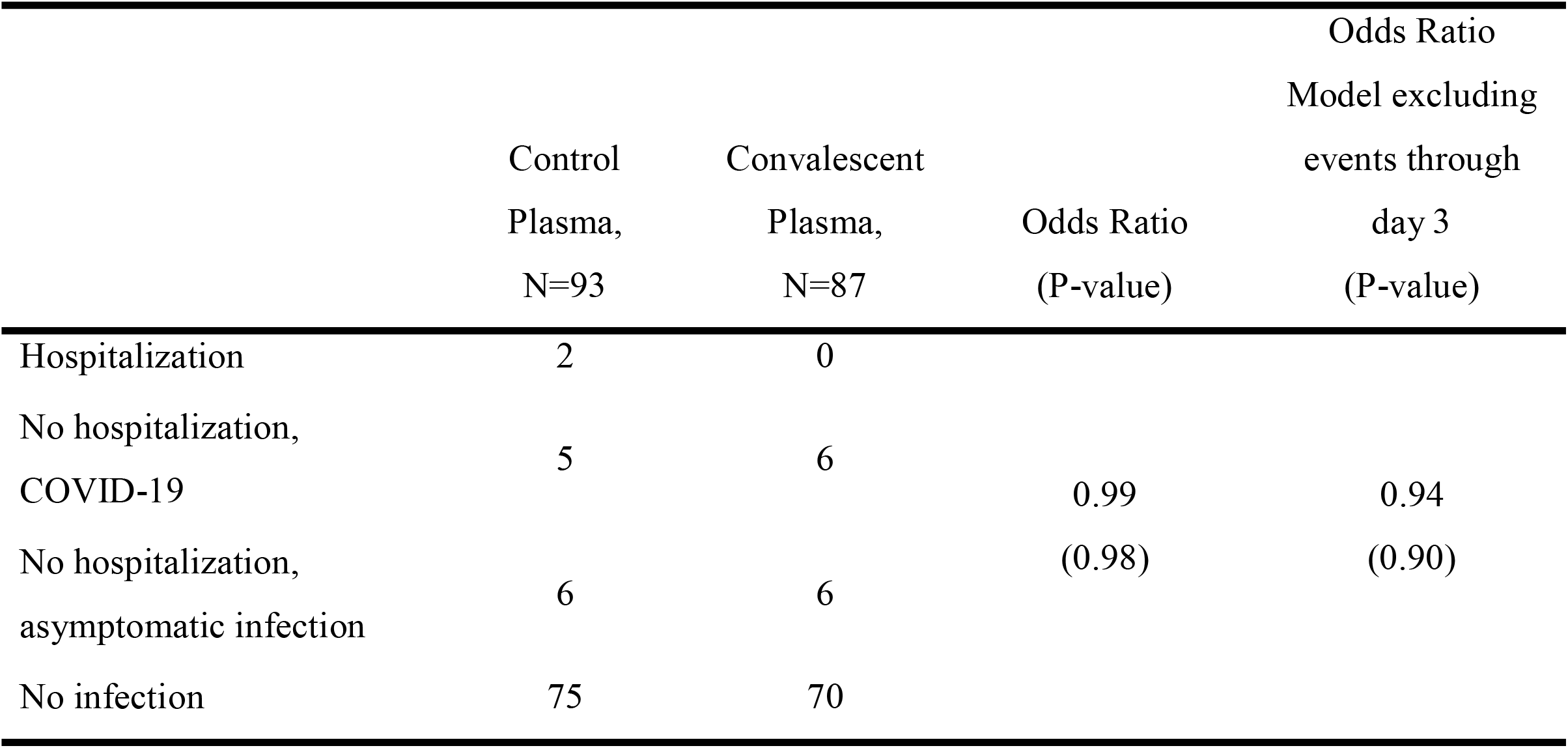
Clinical severity.

|  | Control<br>Plasma,<br>N=93 | Convalescent<br>Plasma,<br>N=87 | Odds Ratio<br>(P-value) | Odds Ratio<br>Model excluding<br>events through<br>day 3<br>(P-value) |
| --- | --- | --- | --- | --- |
| Hospitalization | 2 | 0 |  |  |
| No hospitalization,<br>COVID-19 | 5 | 6 | 0.99<br>(0.98) | 0.94<br>(0.90) |
| No hospitalization,<br>asymptomatic infection | 6 | 6 |  |  |
| No infection | 75 | 70 |  |  |

### Relationship between antibody titers and infection

Anti-S and anti-SRBD IgG AUC titers were not associated with development or time to infection in CCP recipients including when limiting analysis to those developing infection > 3 days after transfusion, Supplemental Material) (Supplemental figure).

## Discussion

This randomized, placebo-controlled, double-blinded trial evaluated the efficacy and safety of a single unit of high antibody titer CCP for prevention of SARS-CoV-2 infection following recent, close contact exposure to a person with COVID-19. In this sample of outpatients exposed to COVID-19, CCP did not reduce SARS-CoV-2 infection in participants transfused up to 120 hours following exposure.

The findings contrast with successful use of mAbs for PEP^8^. Inability of CCP to prevent infection cannot be ascribed to the absence of specific antibody to SARS-CoV-2, as both CCP and the mAbs contain SARS-CoV-2 specific antibodies. Insufficient antibody dose in the CCP used is one explanation for lack of efficacy as PEP. The amount of immunoglobulin in the casirivimab/imdevimab dose is 1.2 grams, likely exceeding the amount of viral-specific antibodies in a unit of high-titer plasma. The concentration of antibodies in the casirivimab and imdevimab PEP trial was 22-25 mg/L which is about 150 times that needed for neutralization of many variants^22,23^. CCP used in this study had neutralizing titers of 1:20 to 1:80, which would be diluted about 20-fold after transfusion, resulting in a 10-100 lower neutralizing capacity than mAbs. Qualitative differences between the products could also affect efficacy. For CCP, much of the neutralizing capacity is in IgM^24^, a large molecule with poor tissue penetration; mAbs are entirely IgG, which has better tissue penetration^25^. Differences in patient populations could also explain the divergent results. The casirivimab/imdevimab PEP study required participants to be household contacts of infected individuals (presumably with close and ongoing exposures). Our study included participants with single close contacts. These differences may have affected our results.

Breakthrough SARS-CoV-2 infections despite vaccination provide insight as to why CCP did not prevent infection. IgG is likely insufficient to prevent upper airways infection, presumably because of insufficient concentration within respiratory airway mucosa during initial infection when the epithelium is intact. As infection progresses an inflammatory response permits transudation of serum (and IgG) into tissues. The large amount of immunoglobulin in plasma after prophylactic mAb administration or vaccination is presumably sufficient to prevent progression of infection. The amount after single-unit CCP administration may be insufficient to affect the course of initial infection, especially if much of the neutralizing antibody is IgM with limited tissue penetration. This is consistent with animal studies reporting antibodies’ inefficiency at reducing virus in nasal tissues^26^. Another possibility is that CCP contained both neutralizing and non-neutralizing antibodies and the latter impaired viral neutralization. Notably, two control participants were hospitalized for COVID-19 (one with hematological disease and hypogammaglobulinemia). Though the numbers are small, none of those who received CCP progressed to hospitalization, thus echoing findings that early treatment reduces progression of disease^11^. CCP may have insufficient specific antibody concentration to prevent infection, but administration of specific antibodies early in infection could avert worse outcomes.

Before the availability of effective vaccines, convalescent serum was used for prophylaxis of measles^27^ and mumps^5^. Although not tested in placebo-controlled randomized trials, serum prevented measles and mumps-related orchitis. Like SARS-CoV-2, these viruses are acquired by the respiratory route, but disease manifestations are systemic^28^. For both measles and mumps, success of convalescent plasma prophylaxis was measured by prevention of systemic disease (rash and orchitis). These experiences suggest that it may be easier to prevent systemic disease with antibodies than against respiratory tract-only infections. A similar pattern is found with pneumococcal vaccine, in which antibodies are more effective in preventing sterile site than respiratory tract disease ^29^.

In this study, CCP was associated with substantially fewer AE’s than control plasma. The reason for this finding is unclear. As there were two COVID-19-related hospitalizations in control recipients and none in CCP, a possible explanation is protection from severe disease in those developing COVID-19 (Sullivan et al; under review). Early in the pandemic, there were concerns about antibody-dependent enhancement (ADE) of infection ^30,31^. While ADE has not been reported in CCP studies to date, almost all were conducted in hospitalized patients^32^ and do not rule out the possibility of ADE in early infection when endogenous antibody responses are lacking. In this study, CCP was administered before or very early in the course of infection and there was no evidence of toxicity or adverse effect. This strongly suggests that ADE is not a significant concern^30,31,33^.

The study had limitations. The logistical challenges were formidable and frequently changed with the evolving pandemic. Enrollment declined precipitously with widespread vaccine availability. Previously vaccinated individuals were ineligible for participation, and guidance to defer vaccination until 90 days after receipt of CCP deterred potential subjects. The enrollment goal of 500 total participants was not achieved. However, conditional power analyses for the primary endpoint of infection suggest that results may not have significantly differed if the trial achieved its target enrollment.

In conclusion, this RCT of high titer CCP given to participants exposed to, but not infected with SARS-CoV-2, within 120 hours demonstrated that CCP was safe. This study did not provide evidence of efficacy and conditional power analysis suggests that a larger sample would not have had a different result. Future studies of CCP prophylaxis might consider a higher dose of antibodies with multiple units or use of higher titer as well as consider targeting populations most at risk including the immunocompromised or elderly, and might consider greater emphasis on clinical rather than laboratory outcomes.

## Data Availability

All data produced in the present study are available upon reasonable request to the authors

## Acknowledgments

The authors gratefully acknowledge the study participants who generously gave of their time and biological specimens. Initiation of this work was catalyzed by grants from Bloomberg Philanthropies and the State of Maryland

Supplemental Figure: Donor and day 1 anti-RBD IgG AUC (left) and titer (right) antibody levels in CCP recipients who remained infection free or developed infection were in the same range. Geomeans are marked with geomean ratio of donor to day one recipient of 40, 14 with AUC and 26 and 14 with titer for not infected (green dots, n=58), and infected (red dots, n=9), respectively.

## References

1. WHO (COVID-19) Homepage. 2021. (Accessed June 12, 2021, 2021, at https://covid19.who.int/.)

2. Coronavirus (COVID-19) Vaccinations. 2021. (Accessed June 13, 2021, 2021, at https://ourworldindata.org/covid-vaccinations.)

3. Boyarsky BJ, Werbel WA, Avery RK, et al. Antibody Response to 2-Dose SARS-CoV-2 mRNA Vaccine Series in Solid Organ Transplant Recipients. JAMA 2021;325:2204–6.

4. Gallagher JR. Use of Convalescent Measles Serum to Control Measles in a Preparatory School. Am J Public Health Nations Health 1935;25:595–8.

5. Rambar AC. MUMPS: Use of Convalescent Serum in the Treatment and Prophylaxis of Orchitis. American Journal of Diseases of Children 1946;71:1–13.

6. Luke TC, Casadevall A, Watowich SJ, Hoffman SL, Beigel JH, Burgess TH. Hark back: passive immunotherapy for influenza and other serious infections. Crit Care Med 2010;38:e66–73.

7. Hemming VG. Use of intravenous immunoglobulins for prophylaxis or treatment of infectious diseases. Clin Diagn Lab Immunol 2001;8:859–63.

8. O’Brien MP, Forleo-Neto E, Musser BJ, et al. Subcutaneous REGEN-COV Antibody Combination to Prevent Covid-19. New England Journal of Medicine 2021;385:1184–95.

9. Cohen MS, Nirula A, Mulligan MJ, et al. Effect of Bamlanivimab vs Placebo on Incidence of COVID-19 Among Residents and Staff of Skilled Nursing and Assisted Living Facilities: A Randomized Clinical Trial. JAMA 2021;326:46–55.

10. Piechotta V, Iannizzi C, Chai KL, et al. Convalescent plasma or hyperimmune immunoglobulin for people with COVID-19: a living systematic review. Cochrane Database Syst Rev 2021;5:Cd013600.

11. Libster R, Pérez Marc G, Wappner D, et al. Early High-Titer Plasma Therapy to Prevent Severe Covid-19 in Older Adults. New England Journal of Medicine 2021.

12. Joyner MJ, Carter RE, Senefeld JW, et al. Convalescent Plasma Antibody Levels and the Risk of Death from Covid-19. New England Journal of Medicine 2021.

13. Thompson MA, Henderson JP, Shah PK, et al. Association of Convalescent Plasma Therapy With Survival in Patients With Hematologic Cancers and COVID-19. JAMA Oncology 2021;7:1167–75.

14. Casadevall A, Henderson JP, Joyner MJ, Pirofski LA. SARS-CoV-2 variants and convalescent plasma: reality, fallacies, and opportunities. J Clin Invest 2021;131.

15. Klein SL, Pekosz A, Park HS, et al. Sex, age, and hospitalization drive antibody responses in a COVID-19 convalescent plasma donor population. J Clin Invest 2020;130:6141–50.

16. Diaz I, Colantuoni E, Hanley DF, Rosenblum M. Improved precision in the analysis of randomized trials with survival outcomes, without assuming proportional hazards. Lifetime Data Anal 2019;25:439–68.

17. Benkeser D, Diaz I, Luedtke A, Segal J, Scharfstein D, Rosenblum M. Improving precision and power in randomized trials for COVID-19 treatments using covariate adjustment, for binary, ordinal, and time-to-event outcomes. Biometrics 2020.

18. Diaz I, Colantuoni E, Rosenblum M. Enhanced precision in the analysis of randomized trials with ordinal outcomes. Biometrics 2016;72:422–31.

19. Royston P, Parmar MK. Flexible parametric proportional-hazards and proportional-odds models for censored survival data, with application to prognostic modelling and estimation of treatment effects. Stat Med 2002;21:2175–97.

20. Royston P. Model selection for univariable fractional polynomials. Stata J 2017;17:619–29.

21. Laud PJ. Equal-tailed confidence intervals for comparison of rates. Pharm Stat 2017;16:334–48.

22. Weinreich DM, Sivapalasingam S, Norton T, et al. REGEN-COV Antibody Combination and Outcomes in Outpatients with Covid-19. N Engl J Med 2021.

23. Copin R, Baum A, Wloga E, et al. The monoclonal antibody combination REGEN-COV protects against SARS-CoV-2 mutational escape in preclinical and human studies. Cell 2021;184:3949–61.e11.

24. Gasser R, Cloutier M, Prévost J, et al. Major role of IgM in the neutralizing activity of convalescent plasma against SARS-CoV-2. Cell Rep 2021;34:108790.

25. Burnett D. Immunoglobulins in the lung. Thorax 1986;41:337–44.

26. Zhou D, Chan JF, Zhou B, et al. Robust SARS-CoV-2 infection in nasal turbinates after treatment with systemic neutralizing antibodies. Cell Host Microbe 2021;29:551–63.e5.

27. Gallagher JR. Use of convalescent measles serum to control measles in a preparatory school. Am J Public Health 1935;25:595–8.

28. Rambar AC. Mumps; use of convalescent serum in the treatment and prophylaxis of orchitis. Am J Dis Child 1946;71:1–13.

29. Webber C, Patton M, Patterson S, Schmoele-Thoma B, Huijts SM, Bonten MJ. Exploratory efficacy endpoints in the Community-Acquired Pneumonia Immunization Trial in Adults (CAPiTA). Vaccine 2017;35:1266–72.

30. Yager EJ. Antibody-dependent enhancement and COVID-19: Moving toward acquittal. Clin Immunol 2020;217:108496.

31. Dzik S. COVID-19 Convalescent Plasma: Now Is the Time for Better Science. Transfus Med Rev 2020;34:141–4.

32. Joyner MJ, Wright RS, Fairweather D, et al. Early safety indicators of COVID-19 convalescent plasma in 5000 patients. The Journal of clinical investigation 2020;130:4791–7.

33. Joyner M, Bruno K, Stephen A. Klassen S, et al. Safety Update: COVID-19 Convalescent Plasma in 20,000 Hospitalized Patients. Mayo Clin Proc 2020.

