## Supplemental material for "Randomized controlled trial transfusing convalescent plasma as post-exposure prophylaxis against SARS-CoV-2 infection"

Appendix: Supplemental Material

Contents:

I. Table of grade 3 or 4 Adverse Events
II. Listing of Serious Adverse Events
III. Antibody AUC models
IV. Statistical Analysis Plan

### I. **Cumulative incidence of grade 3 or 4 adverse events by treatment status**

(As of 06 July 2021)

|  | **Control** | **CCP** |
| --- | --- | --- |
| **Grade 3 or 4 events** | **13** | **4** |
| **Total person-years** | 18.64 | 17.05 |
| Rate per person-years | 0.70 cases/py | 0.23 cases/py |
| Median [IQR} observation time, days | 90 [88, 92] | 90 [88, 91] |
| Rate difference (95% CI) |  | 0.46 (0.02, 0.91) |
| **abdominal pain** | **1** | **0** |
| **Total person-years** | 18.64 | 17.05 |
| Rate per person-years | 0.05 cases/py | 0.00 cases/py |
| Median [IQR} observation time, days | 90 [88, 92] | 90 [88, 91] |
| Rate difference (95% CI) |  | 0.05 (-0.05, 0.15) |
| **atrial fibrillation** | **1** | **0** |
| **Total person-years** | 18.64 | 17.05 |
| Rate per person-years | 0.05 cases/py | 0.00 cases/py |
| Median [IQR} observation time, days | 90 [88, 92] | 90 [88, 91] |
| Rate difference (95% CI) |  | 0.05 (-0.05, 0.15) |
| **generalized muscle weakness** | **1** | **0** |
| **Total person-years** | 18.64 | 17.05 |
| Rate per person-years | 0.05 cases/py | 0.00 cases/py |
| Median [IQR} observation time, days | 90 [88, 92] | 90 [88, 91] |
| Rate difference (95% CI) |  | 0.05 (-0.05, 0.15) |
| **hypertension** | **2** | **0** |
| **Total person-years** | 18.64 | 17.05 |
| Rate per person-years | 0.11 cases/py | 0.00 cases/py |
| Median [IQR} observation time, days | 90 [88, 92] | 90 [88, 91] |
| Rate difference (95% CI) |  | 0.11 (-0.04, 0.26) |
| **hypoxia** | **1** | **0** |
| **Total person-years** | 18.64 | 17.05 |
| Rate per person-years | 0.05 cases/py | 0.00 cases/py |
| Median [IQR} observation time, days | 90 [88, 92] | 90 [88, 91] |
| Rate difference (95% CI) |  | 0.05 (-0.05, 0.15) |
| **Transfusion reactions** | **1** | **0** |
| **Total person-years** | 18.64 | 17.05 |
| Rate per person-years | 0.05 cases/py | 0.00 cases/py |
| Median [IQR} observation time, days | 90 [88, 92] | 90 [88, 91] |
| Rate difference (95% CI) |  | 0.05 (-0.05, 0.15) |

|  | **Control** | **CCP** |
| --- | --- | --- |
| **neutrophil count <500** | **1** | **0** |
| **Total person-years** | 18.64 | 17.05 |
| Rate per person-years | 0.05 cases/py | 0.00 cases/py |
| Median [IQR} observation time, days | 90 [88, 92] | 90 [88, 91] |
| Rate difference (95% CI) |  | 0.05 (-0.05, 0.15) |
| **Pneumonia (any)** | **3** | **0** |
| **Total person-years** | 18.64 | 17.05 |
| Rate per person-years | 0.16 cases/py | 0.00 cases/py |
| Median [IQR} observation time, days | 90 [88, 92] | 90 [88, 91] |
| Rate difference (95% CI) |  | 0.16 (-0.02, 0.34) |
| **sepsis** | **1** | **0** |
| **Total person-years** | 18.64 | 17.05 |
| Rate per person-years | 0.05 cases/py | 0.00 cases/py |
| Median [IQR} observation time, days | 90 [88, 92] | 90 [88, 91] |
| Rate difference (95% CI) |  | 0.05 (-0.05, 0.15) |
| **cardiac disorders - Other** | **0** | **1** |
| **Total person-years** | 18.64 | 17.05 |
| Rate per person-years | 0.00 cases/py | 0.06 cases/py |
| Median [IQR} observation time, days | 90 [88, 92] | 90 [88, 91] |
| Rate difference (95% CI) |  | -0.06 (-0.17, 0.06) |
| **chest wall pain** | **0** | **1** |
| **Total person-years** | 18.64 | 17.05 |
| Rate per person-years | 0.00 cases/py | 0.06 cases/py |
| Median [IQR} observation time, days | 90 [88, 92] | 90 [88, 91] |
| Rate difference (95% CI) |  | -0.06 (-0.17, 0.06) |
| **hyponatremia** | **0** | **1** |
| **Total person-years** | 18.64 | 17.05 |
| Rate per person-years | 0.00 cases/py | 0.06 cases/py |
| Median [IQR} observation time, days | 90 [88, 92] | 90 [88, 91] |
| Rate difference (95% CI) |  | -0.06 (-0.17, 0.06) |
| **vasovagal reaction** | **0** | **1** |
| **Total person-years** | 18.64 | 17.05 |
| Rate per person-years | 0.00 cases/py | 0.06 cases/py |
| Median [IQR} observation time, days | 90 [88, 92] | 90 [88, 91] |
| Rate difference (95% CI) |  | -0.06 (-0.17, 0.06) |

II. **Listing of Serious Adverse Events**

| **SR#** | **Treatment** | **Age range (years)** | **Gender** | **SAE description and grade** | **Reason reported** | **Outcome** |
| --- | --- | --- | --- | --- | --- | --- |
| 1 | Control Plasma | 60-69 | F | Blood and lymphatic system disorders - Other, specify^1^; grade 3 | Hospitalization | Resolved |
| 2 | Control Plasma | 60-69 | F | Sepsis; grade 3 | Hospitalization | Resolved |
| 3 | Control Plasma | 50-59 | F | COVID-19 Pneumonia; grade 3 | Hospitalization | Resolved |
| 4 | Control Plasma | 20-29 | M | Infusion related reaction^2^; grade 3 | Significant medical event | Resolved |
| 5 | Control Plasma | Over 80 | M | Pneumonia; grade 3 | Hospitalization | Resolved |
| 6 | Control Plasma | 60-70 | M | Atrial fibrillation and COVID-19 pneumonia | Hospitalization | Resolved |

^1^Febrile neutropenia

^2^Transfusion-related reaction

Total number of SAEs is 6 in 5 subjects

**Supplemental Figure Legend:** Donor and day 1 anti-RBD IgG AUC (left) and titer (right) antibody levels in CCP recipients who remained infection free or developed infection were in the same range. Geomeans are marked with geomean ratio of donor to day one recipient of 40, 14 with AUC and 26 and 14 with titer for not infected (green dots, n=58), and infected (red dots, n=9), respectively.

III. **Antibody AUC Models**

Primary outcome of any infection after transfusion

A fractional polynomial model with a flexible Weibull survival model was used to estimate the hazard ratio (HR) for *any infection after transfusion*. Both spline and fractional polynomial models were assessed with the lowest Akiake’s information criterion used to choose the final model.

For the anti-spike antibody area under the curve (AUC), the final model was a fractional polynomial model of the first degree with AUC levels raised to the 3^rd^ power. The likelihood ratio test for inclusion of anti-spike AUC provided a p-value of 0.11. The following table provides the hazard ratio (HR) at different quantiles of the anti-spike AUC level.

| **Hazard ratio for progression to any infection by day 28 visit after transfusion by quantile of anti-spike AUC as estimated from a flexible Weibull time to event model with fractional polynomial to allow for non-linearity** | | | | |
| --- | --- | --- | --- | --- |
| **Quantile** | **Anti-spike AUC** | **HR** | **Standard Error** | **One-sided 95% Confidence Interval** |
| **0.10** | 1670 | 1.00 | 0.000068 | 1.00 |
| **0.30** | 3236 | 1.00 | 0.00041 | 1.00 |
| **0.50** | 5816 | 1.00 | 0.0021 | 1.00 |
| **0.70** | 10972 | 0.99 | 0.013 | 1.02 |
| **0.90** | 20680 | 0.96 | 0.087 | 1.11 |

The final model for anti-spike receptor binding domain (SRBD) AUC was a fractional polynomial with first degree where the AUC levels were raised to 0.5 power. The likelihood ratio test provided a p-value of 0.12. The following table provides the hazard ratio (HR) at different quantiles of the anti-SRBD AUC level.

| **Hazard ratio for progression to any infection by day 28 visit after transfusion by quantile of anti-SRBD AUC as estimated from a flexible Weibull time to event model with fractional polynomial to allow for non-linearity** | | | | |
| --- | --- | --- | --- | --- |
| **Quantile** | **Anti-spike AUC** | **HR** | **Standard Error** | **One-sided 95% Confidence Interval** |
| **0.10** | 658 | 1.03 | 0.08 | 1.18 |
| **0.30** | 1399 | 1.05 | 0.14 | 1.31 |
| **0.50** | 2637 | 1.08 | 0.20 | 1.49 |
| **0.70** | 5016 | 1.11 | 0.29 | 1.80 |
| **0.90** | 13494 | 1.20 | 0.51 | 2.78 |

Pre-specified sensitivity analysis restricting events to after 3 days following transfusion

A fractional polynomial model with a flexible Weibull survival model was used to estimate the hazard ratio (HR) for *any infection occurring after 3 days following transfusion*. Both spline and fractional polynomial models were assessed with the lowest Akiake’s information criterion used to choose the final model.

For the anti-spike antibody area under the curve (AUC), the final model was a fractional polynomial model of the first degree with AUC levels raised to the 2^nd^ power. The likelihood ratio test for inclusion of anti-spike AUC provided a p-value of 0.11. The following table provides the hazard ratio (HR) at different quantiles of the anti-spike AUC level.

| **Hazard ratio for progression to any infection by day 28 visit after transfusion by quantile of anti-spike AUC as estimated from a flexible Weibull time to event model with fractional polynomial to allow for non-linearity** | | | | |
| --- | --- | --- | --- | --- |
| **Quantile** | **Anti-spike AUC** | **HR** | **Standard Error** | **One-sided 95% Confidence Interval** |
| **0.10** | 1674 | 0.98 | 0.004 | 0.99 |
| **0.30** | 3212 | 0.94 | 0.02 | 0.98 |
| **0.50** | 5816 | 0.83 | 0.12 | 1.01 |
| **0.70** | 10734 | 0.55 | 0.71 | 1.76 |
| **0.90** | 20680 | 0.11 | 4.91 | 359 |

The final model for anti-spike receptor binding domain (SRBD) AUC was a fractional polynomial with first degree where the AUC levels were raised to 3^rd^ power. The likelihood ratio test provided a p-value of 0.10. The following table provides the hazard ratio (HR) at different quantiles of the anti-SRBD AUC level.

| **Hazard ratio for progression to any infection by day 28 visit after transfusion by quantile of anti-SRBD AUC as estimated from a flexible Weibull time to event model with fractional polynomial to allow for non-linearity** | | | | |
| --- | --- | --- | --- | --- |
| **Quantile** | **Anti-spike AUC** | **HR** | **Standard Error** | **One-sided 95% Confidence Interval** |
| **0.10** | 658 | 1.00 | 0.0002 | 1.00 |
| **0.30** | 1399 | 1.00 | 0.002 | 1.00 |
| **0.50** | 2637 | 0.99 | 0.009 | 1.01 |
| **0.70** | 4658 | 0.97 | 0.05 | 1.05 |
| **0.90** | 12065 | 0.63 | 0.81 | 2.42 |

IV. **Statistical Analysis Plan**

The following is the statistical analysis plan that was agreed upon between the data coordinating center and principal investigator prior to analyses being conducted on final data.

**Convalescent Plasma to Stem Coronavirus: A Randomized Controlled Double Blinded Phase 2 Study Comparing the Efficacy and Safety of Human Coronavirus Immune Plasma (HCIP) vs. Control (SARS-CoV-2 non-immune plasma) among Adults Exposed to COVID-19**

**NCT** **04323800**

**Statistical Analysis Plan Version 2.1**

**Based on Protocol Version December 23, 2020**

**May 12, 2021**

**Document History**

**Version 1.0 (28 July 2020);** Document developed from Protocol version dated June 9, 2020

**Version 2.0 (26 April 2021);** Document developed from Protocol version dated December 23, 2020

**Version 2.1 (12 May 2021);** Document developed from Protocol version dated December 23, 2020

**Roles and Responsibilities**

This statistical analysis plan (SAP) for CSSC-001 study has been reviewed by the PI, senior trial statisticians and DCC leads.

**Introduction**

There are currently few proven treatment options for coronavirus disease (COVID-19), which is caused by Severe Acute Respiratory Syndrome Coronavirus 2 (SARS-CoV-2). Human convalescent plasma has been successfully used for prevention and treatment of other infections and thus may provide an option for treatment of COVID-19 and could be rapidly available from people who have recovered from disease and can donate plasma.

Passive antibody therapy involves the administration of antibodies against a given infectious agent to a susceptible or ill individual for the purpose of treating an infectious disease caused by that agent. In contrast, active vaccination requires the induction of an immune response to the vaccine that takes time to develop and varies depending on the vaccine recipient. Some immunocompromised patients fail to achieve an adequate immune response. Thus, passive antibody administration, in some instances, represents the only means of providing immediate immunity to susceptible persons and more predicable immunity for highly immunocompromised patients.

A general principle of passive antibody therapy is that it is more effective when used for prophylaxis than for treatment of disease. When used for therapy, antibody is most effective when administered shortly after the onset of symptoms. The reason for temporal variation in efficacy is not well understood but could reflect that passive antibody works by neutralizing the initial inoculum, which is likely to be much smaller load of infection than that once the disease is established. Another explanation is that antibody works by modifying the inflammatory response, which is also easier during the initial immune response, which may be asymptomatic [Casadevall A, and Pirofski LA. Antibody-mediated regulation of cellular immunity and the inflammatory response. *Trends Immunol.* 2003;24(9):474-8]. As an example, passive antibody therapy for pneumococcal pneumonia was most effective when administered shortly after the onset of symptoms and there was no benefit if antibody administration was delayed past the third day of disease [Casadevall A, and Scharff MD. "Serum Therapy" revisited: Animal models of infection and the development of passive antibody therapy. *Antimicrob Agents Chemotherap.* 1994;38(1695-702)].

In CSSC-001, we hypothesize that convalescent plasma will reduce the cumulative incidence of COVID-19 disease among individuals who have been exposed to COVID-19 but are not known to be infected. A multi-center randomized clinical trial will be conducted to test this hypothesis. Our primary hypothesis is that by providing anti-SARS-CoV-2 plasma, the cumulative incidence of COVID-19 disease, defined as symptoms compatible with infection and RT-PCR positive for SARS-CoV-2, is less than the cumulative incidence in individuals randomized to receive transfusion with SARS-CoV-2 non-immune plasma. A secondary hypothesis is that individuals receiving anti-SARS-CoV-2 convalescent plasma are likely to have less disease severity than a control group.

**Study Design**

CSSC-001 is a two-arm, parallel-group, multi-center, randomized superiority trial in which outpatient adults with recent exposure to SARS-CoV-2 but no current infection will be randomized to receive either human coronavirus immune plasma (HCIP) or control (SARS-CoV-2 non-immune) plasma. This randomized, double-blind, controlled, phase 2 trial will assess the efficacy and safety of HCIP to reduce the cumulative incidence of the development of COVID-19 during study follow-up. Adults 18 years of age or older, with high risk exposure (see below for definition) to person with COVID-19 within 96 hours of enrollment (and 120 hours of receipt of plasma). A total of approximately 500 eligible subjects will be randomized in a 1:1 ratio to either HCIP or control plasma.

**High risk exposure will be defined** as currently defined by CDC and modified as the CDC makes changes. Currently this includes any close contact exposure (as defined by CDC guidelines) to person with COVID-19. Health Care Personnel (HCP) are defined as physicians, nurses, nursing assistants, therapists, technicians, emergency medical service personnel, dental personnel, pharmacists, laboratory personnel, autopsy personnel, students, trainees and contractual staff. Persons not directly involved in patient care such as clerical, dietary, housekeeping, laundry, security, maintenance, billing, and volunteers, but who are potentially exposed to SARS-CoV-2 in the healthcare setting, also fall under the category of HCP in this protocol.

*Primary Outcome:*

- The primary outcome of CSSC-001 will be the cumulative incidence by day 28 of the development of COVID-19 (symptoms compatible with infection and/or RT-PCR positive) regardless of disease severity comparing those randomized to HCIP to those who are transfused with control plasma.

*Secondary Outcome:*

- Compare the cumulative incidence of disease severity between those randomized to HCIP and control plasma. Severity of disease will be measured using a clinical event scale of disease severity evaluated up to Day 28:

1. Death

2. Requiring mechanical ventilation and/or in ICU

3. non-ICU hospitalization, requiring supplemental oxygen;

4. non-ICU hospitalization, not requiring supplemental oxygen, or a stay of >24 hours for observation in an ED, field hospital, or other healthcare unit*, or any receipt of O2 for >24 hours, outside of hospital*

5. Not hospitalized, but with clinical and laboratory evidence^[1]^ of COVID-19 infection (symptomatic infection)

6. Not hospitalized, no clinical symptoms compatible with COVID-19, but with laboratory evidence of COVID-19 infection (asymptomatic infection)

7. No symptoms compatible with infection, no laboratory evidence of COVID-19 (No indication of infection)

*Other Outcomes*:

- Comparison of anti-SARS-CoV-2 titers at days -1 to 0, 1, 7 ,14, and 90 days after transfusion between those randomized to HCIP and control plasma
- Comparison of rates and durations of SARS-CoV-2 PCR positivity (RT PCR) at days -1 to 0, 1,7,14, and 28 between treatment groups
- Comparison of levels of SARS-CoV-2 RNA amongst those randomized to HCIP vs. control plasma at days -1 to 0, 1, 7, 14, and 28

**Power and Sample Size**

We hypothesize that outpatient adults with recent exposure to COVID-19 who receive HCIP will have a lower incidence of developing a SARS-CoV-2 infection with COVID-19 when compared with similar adults receiving control plasma. A sample size of 500 individuals with a high risk exposure within 96 hours of enrollment with a 1:1 allocation ratio to each treatment group will have 80% power to detect a 50% reduction in cumulative incidence by 28 days. This is with a one-sided type 1 error of 5% as we are solely interested in superiority. We assumed that the probability of incident COVID-19 cases is between 0.10 and 0.20 in the control population. We explored several different scenarios with an exponential model to identify the lambda parameter and the package ‘powerSurvEpi’ for the R statistical software was then used to calculate the sample sizes for these scenarios (See Table1). We assumed that the cumulative incidence of symptomatic disease in SARS-CoV-2 exposed individuals randomized to control plasma to be 0.13X by day 28 and 0.065 (50% reduction in risk) by day 28 among those randomized to HCIP. We therefore estimated 488 participants (244 in each arm). It is anticipated that very few of these subjects will be randomized and not start study plasma infusion (and so be excluded from the primary analysis). Furthermore, it is anticipated that there will be low loss to follow-up prior to development of disease or 28 days (and so have missing data for the primary endpoint). Therefore, we have extended the total sample size to 500 participants. For all analyses, we will use a modified intention-to-treat analyses, which excludes randomized subjects who do not initiate an infusion of the study plasma.

| Table 1: Total sample size by the incident proportion of COVID-19 among controls and by percent risk reduction | | |
| --- | --- | --- |
| Proportion of Incident COVID-19 cases at day 28 among controls | 50% Risk Reduction | 75% Risk Reduction |
| 0.10 | 648 | 199 |
| 0.13 | **488** | 150 |
| 0.15 | 415 | 129 |
| 0.20 | 298 | 94 |

**Randomization**

The Data Coordinating Center (DCC) will work with the EDC developers (Prelude Dynamics) to generate random treatment assignments using a documented process. The randomization schedule will be designed to yield an expected allocation ratio of 1:1 for HCP + control plasma. Assignments will be stratified by clinical site and schedules will employ permuted block designs, with block sizes to be determined and documented at the DCC. Adjustment for residual or other imbalances in the baseline composition of the treatment groups, if needed, will be done using multiple regression techniques rather than through further stratification in the design.

Treatment assignments will be masked to the participants and the personnel of the clinical sites, but not to an unmasked statistician associated with the DCC. This unmasked statistician will only provide unmasking information (1) to the DSMB, (2) if required for participant safety, and (3) to the analytical team after databases have been locked. Unforeseen circumstances that may require unmasking will need to be approved by the Steering Committee. Staff in the blood banks will also be unmasked to treatment assignments

Treatments will be assigned at the baseline visit using an online program accessible to the clinical sites. After the entry of specified pre-randomization data, each enrolled participant’s ID will be irrevocably linked to the next unassigned treatment for that clinical site. Upon notification of a randomization, an unmasked email is sent to the blood bank informing of the participant identifier and the treatment group assignment. Blood bank personnel will select an appropriate unit of plasma for transfusion and will mask (over-label) the unit consistent with local and federal regulations. The data system will also check for and prevent duplicate assignments (same participant randomized more than once). The treatment assignment tables in the data system will be encrypted to prevent inadvertent disclosures.

The procedures related to randomization of participants at the clinical sites will be as follows:

- Clinical sites will collect randomization eligibility and baseline data on the appropriate data collection instruments and will enter these data into the database.
- The data system will confirm randomization eligibility, issue the next assignment, and will relay treatment assignments to the DCC (masked) and blood bank (unmasked) as described above.
- The data system will automatically store the date and time of assignment, the identity of the clinical site personnel making the assignment, the participant’s ID, and the treatment group assignment.

The data system will provide access to randomization materials, including a visit schedule and allowable time windows for visits.

**Statistical Principles**

General principles for analysis include the following:

- The primary analysis will be performed according to the participants’ original randomized treatment groups excluding those who do not initiate transfusion of study plasma (modified intention to treat).
- All participants, including those who have withdrawn from the study or were found to be ineligible after randomization, will be included in their assigned treatment group and analyzed for safety.
- All outcomes, including death, following randomization will be included. Death in the absence of known infection will be presumed to be indicative of infection.
- Missing data will be minimized by study design and conduct. It will be addressed analytically using multiple imputation methods

Analyses will be done to explore differences in the outcomes between the treatment groups. Results of these analyses will be presented unadjusted (crude) and adjusted for covariates. Variables chosen for adjustment are specifically for increasing precision in estimates of treatment efficacy and thus must be predictive of disease outcome [Diaz et al, Lifetime Data Anal (2019): 25]. To identify the adjustment variables, we will utilize a hybrid approach of pre-specifying some variables and using an algorithmic approach to identify variables to adjust for among pre-specified candidate variables. Variables that we are near certain to be predictive of outcome will be adjusted for. Age has been consistently related to worse outcomes for COVID-19 and therefore will be included in analyses for adjustment. Other pre-specified variables that will be candidates for inclusion in primary analysis will be determined via an algorithmic approach and these variables will include: clinical site, race, ethnicity, sex, category of exposure, hematology factors and other laboratory markers (i.e., CBC and metabolic panels), body mass index, ABO blood group, prescription steroid, targeted physical exam, and prior comorbidities that have specifically been associated with worse COVID-19 outcomes including: asthma, chronic kidney disease, chronic lung disease (COPD, idiopathic pulmonary fibrosis, cystic fibrosis), diabetes, hemoglobin disorders (thalassemia, sickle cell disease), immunocompromising conditions (cancer, HIV, organ transplantation, prolonged use of corticosteroids), chronic liver disease, hypertension, and serious heart conditions (heart failure, coronary artery disease, cardiomyopathies, pulmonary hypertension), obesity, smoking status, dementia, down syndrome, pregnancy, stroke/cerebrovascular disease, and substance use disorders [updated to CDC list on website on 5/29/2021 and Guan et al Eur Respir J. 2020 May; 55(5)]. To determine which of these pre-specified candidate variables to be included, we will conduct variable selection by random survival forest in the entire sample (i.e., not including an indicator term for treatment arm) and blinded to treatment allocation. The variable importance and 95% confidence intervals [Ishwaran et al Statistics in Medicine. 2019;38] shall be used to identify predictive variables for the outcome and included in analytical models. Specifically, variables in which the 95% CI for the variable importance from the random forest does not contain 0 will be adjusted for. This should reduce the number of variables that the analysis adjusts for in order to minimize the degrees of freedom that are used while allowing the analysis to include the variables that have the most correlation with the outcome in order to maximize precision. This hybrid approach will be done on the full sample (i.e., modified ITT sample) and not include the treatment arm (i.e., among entire sample without controlling for convalescent or control plasma) in order to identify the prognostic baseline variables for entire sample.

The pre-randomization variables listed above (age and the pre-specified candidate variables) will be explored and described according to summary statistics (mean and variance or 25^th^, 50^th^, and 75^th^ percentiles or proportion as appropriate for type of variable) by treatment group and overall. Analyses will be done in the R statistical software.

There will be two formal reviews of interim results. The first will be after 15 infections which is expected to occur near 150 participants reaching day 28 and the second after 50% of the total recruitment has been achieved and these individuals followed to at least day 28. Interim analyses will only be adjusted for age. Stopping guidelines will be based on O’Brien-Flemming boundary. The interim analysis Z-value boundary of 3.03 (nominal p-value of 0.0011, spent alpha 0.0011), 2.38 (0.0087, 0.008), and 1.68 (0.464, 0.408) for a one-sided test with Type 1 of 0.05.

**Primary Analysis:**

Our primary hypothesis is that by providing anti-SARS-CoV-2 plasma, the cumulative incidence of the development of active infection will be lower among the individuals receiving HCIP as compared to those receiving control plasma over the course of follow-up. Therefore, our primary endpoint is the development of COVID-19 within the follow-up period. For clarity the working definition of our endpoint is positive for SARS-CoV-2 by molecular test with or without any symptoms compatible with COVID-19. To allow to differentiate between all infections and asymptomatic infections, the analyses will be done using 1) all infections as indicated by research based RT-PCR or capture of positivity by clinical testing and then again for 2) symptomatic infection based upon symptoms compatible with COVID-19 infection and being positive by molecular test (research sample or clinical result). For the primary analysis, individuals who test positive by a molecular test on the calendar day of transfusion will be excluded from the analysis. Individuals whose specimens obtained at screening visit who test positive on two antibody tests done by Dr. Oliver Laeyendecker’s laboratory using the Euroimmune assay and Dr. Sabra Klein using antibody ELISA end point titer and area under curve, will be excluded as indication of a prior unknown COVID-19 infection.

Our analysis will be a time to event analysis examining the effect of anti-SARS-CoV-2 plasma. We will estimate the survival function for each treatment arm in order to estimate the risk difference over time as well as the restricted mean survival time which is the area under the survival function and provides the expected mean time to infection up to time *t*^22^. Our approach will be to estimate the cumulative incidence using the doubly robust estimator based upon targeted minimum loss (TMLE) based estimator as described by Diaz et al (2019). By adjusting for baseline covariates that are related to the outcome, we increase precision. This TMLE based approach was shown to increase precision by around 10% to 20% over an inverse probability weighted or augmented inverse probability weighted estimator [Diaz 2019]. As stated above, we will use a hybrid approach of adjusting for pre-specified variables (age) and an algorithmic approach to identify additional variables related to the outcome. Specifically, a random survival forest will be used to identify variables that are related to the outcome in order to increase precision. We will use a random survival forest blinded to the treatment arm allocation and *not* including an indicator variable for treatment (pre-randomization variables to be considered are listed above).

The primary outcome analysis will be the adjusted comparison of proportions of cumulative incidence of SARS-CoV-2 infection (as indicated by RT-PCR positive) at day 28 comparing the risk-difference and restricted mean survival time between treatment groups with confidence intervals and using a significance threshold of p=0.047 (p-value adjusted for interim analysis) for a one-sided test. This will be repeated for analysis of the cumulative incidence of SARS-CoV-2 symptomatic disease as defined as having a confirmed SARS-CoV-2 positive test and at least one symptom including new/incident: fever, cough, sore throat, malaise, headache, muscle pain, nausea, vomiting, loss of taste, loss of smell, shortness of breath, or dyspnea as per the FDA recommendations in Appendix A within “COVID-19: Developing Drugs and Biological Products for Treatment or Prevention: Guidance for Industry [<https://www.fda.gov/media/137926/download>]. Therefore, we will be assessing both the effect of convalescent plasma on overall infection including asymptomatic disease and its effect on symptomatic disease.

Additional sensitivity analysis include:

1. A per-protocol analysis that accounts for those who received a transfusion and were not transfused with their specific treatment group (i.e., non-adherence) using inverse probability weights to account for non-adherence.

2. Classifying participants with missing outcome data at either day 14 or day 28 as having developed COVID-19 with date of COVID-19 being drawn from distribution reflecting the corresponding treatment arms;

3. Multiple imputation of the development of COVID-19 for missing outcomes;

4. A complete case analysis;

5. As convalescent plasma is unlikely to have an immediate effect after transfusion, we will exclude any events occurring between transfusion and 3 calendar days afterwards;

6. As symptoms compatible with COVID-19 may be non-specific and increase false positives, we had elected to use molecular test with or without COVID-19 symptoms as our outcome for any infection and positive test with at least 1 symptom compatible with COVID-19 to define symptomatic infection. As a sensitivity analysis we will conduct a series of analyses that iteratively increases the number of symptoms compatible with COVID-19 required to be present to define symptomatic COVID-19 infection (e.g., ≥2, ≥3, …). Any individuals who have a pre-existing symptom (perhaps due to prior co-morbidity) that maximizes the number of incident symptoms will be considered positive for any of the sensitivity analyses that are above what they can have. .

**Secondary Analyses:**

A secondary hypothesis is that individuals receiving anti-SARS-CoV-2 convalescent plasma are likely to have less disease severity than control. We have devised a clinical event scale for disease severity based on the COVID clinical severity scale which is composed of clinical events that range from being not infected to death. The event scale is as follows:

1. Death

2. Requiring mechanical ventilation and/or in ICU

3. non-ICU hospitalization, requiring supplemental oxygen;

4. non-ICU hospitalization, not requiring supplemental oxygen

or

a stay of >24 hours for observation in an ED, field hospital, or other healthcare unit*

or

any receipt of O2 for >24 hours, outside of hospital*

5. Not hospitalized, but with clinical and laboratory evidence^[2]^ of COVID-19 infection (symptomatic infection)

6. Not hospitalized, no clinical symptoms compatible with COVID-19, but with laboratory evidence of COVID-19 infection (asymptomatic infection)

7. No symptoms compatible with infection, no laboratory evidence of COVID-19 (No indication of infection)

This analysis will be done using the status of participants by day 32 (corresponding to day 28 visit allowing for window around the scheduled visit) using a doubly robust estimators for ordinal outcomes and adjusted for the pre-specified candidate variables selected via the algorithm as described above. The doubly robust estimator that will be used is that as outlined by Benkeser et al. (2020) based upon a non-parametric extension of the log-odds ratio by Diaz et al. (2016). A sensitivity analysis will be done in which participants who have indication of positive infection (e.g. clinical scale event <7) at ≤3 days after transfusion.

**Other Analyses:**

Other analyses for this study are to determine i) the anti-SARS-CoV-2 antibody titer for individuals at days -1 to 0, 1, 7, 14, and 28 days post-randomization, ii) compare rates and duration of SARS-CoV-2 RNA positivity, and iii) compare levels of SARS-CoV-2 RNA.

Analysis of titers will also primarily be descriptive, comparing the geometric mean titers at days 0, 1, 7, 14, and 90 between the randomized arms. Furthermore, it is of interest to describe the entire distributions of anti-SARS-CoV-2 titers by randomized arms and contrast these distributions. Therefore, we will use quantile regression in order to describe whether there is a shift or change in the titer distribution between randomized arms [Koenker, Quantile Regression, 2005]. Quantile regression does not require the assumption of a parametric or any other type of distribution as it identifies the titer at each percentile. Given that this is a repeated measurement at days 0, 1, 7, 14, and 90 we will account for the correlation within individuals using a cluster bootstrap in order to properly estimate the p-value and 95% confidence intervals.

Analysis of the rate and duration of SARS-CoV-2 PCR positivity between the randomized arms will primarily be descriptive examining proportion positive at days 0, 1, 7, 14, and 28 and then among those who are positive whether individuals lose positivity status at a subsequent visit. To determine the proportion that are positive at each visit, we will do a pooled complementary log-log model in order to describe the cumulative incidence of SARS-CoV-2 PCR positivity over time. The pooled complementary log-log model is a discrete time-to-event-analysis that estimates the log hazard rate at each discrete time point. From this, a cumulative incidence of positivity can be estimated. To determine the duration of positivity, the analysis is complicated by the exact day that an individual becomes positive and the exact day that an individual becomes negative is not known since SARS-CoV-2 PCR positivity will only be acquired at days 0, 1, 7, 14, and 28. However, we can estimate a minimum and maximum amount of time that an individual was positive. For instance if an individual is first positive visit at day 1 and then is positive at day 7 but negative at day 14, then we know that this individual became positive between day 0 and 1 and negative between day 7 and 14. Therefore, the minimum amount of time positive is 7 days (day 8 – day 1) and the maximum is 14 days (day 14 – day 0). Therefore, we can interval censor these individuals. That is, we know that the duration is between 7 and 14 days for this example individual. Across all individuals we can describe the duration of positivity either using a non-parametric approach for time-to-event analysis, but more likely given the sample size, a parametric model. We will assess several parametric distributions aiming for parsimony in the number of parameters being estimated due to the interval censored data which results in increased uncertainty in the model. To determine the best model, we will use Akaike’s Information Criterion (AIC) to choose the best model fit. However, if the sample that becomes positive is small, then we will only be able to describe the observations without a formal statistical model.

We will also describe the distribution of SARS-CoV-2 RNA between randomized arms. This is similar in objective to the comparison of anti-SARS-CoV-2 titers. Therefore, we will use the same approach as above of applying quantile regression.

**Handling of Missing Data:**

Every effort will be made to minimize missing data in the trial. For participants with missing covariate or outcome data, multiple imputation will be used for sensitivity analyses. Specifically, chained equations (MICE) will be used to predict missing variables but also to generate uncertainty around imputed point estimates.

We will employ recommended strategies to prevent missing data:

- We have developed a data collection schedule that will minimize participant burden and we will follow participants according to the data collection schedule regardless of compliance with the study intervention;
- We will maintain frequent contact with the participants through visit reminder calls, texts, or emails;
- We will provide a 24-hour phone number, which participants can contact for questions and support;
- We will employ rigorous training of clinic staff emphasizing the importance of:
  - Congenial interpersonal relationships between the participants and study staff;
  - Using the consent process to ensure that potential participants understand the commitment that they are making;
  - Addressing concerns if participants are dissatisfied so that the participant will remain in the trial;
  - Attempting to collect as much data as possible even if a participant cannot come to the clinic because of other obligations.

We will collect data on timing and reasons for study dropout by treatment group to present in the CONSORT diagram. If participants who drop out of the study appear to be doing so for reasons related to the study, i.e., not missing completely at random, we will perform sensitivity analyses using methods that have been previously described^26^, such as multiple imputation techniques, best- and worst-case scenarios, and correlates of the drop out event included in the models. There are no standard statistical techniques for dealing with data that are missing not at random (MNAR). We will explore one or more of the sensitivity analyses for MNAR given in the NRC report [National Research Council, 2010]. Baseline characteristics of participants with missing measures will be compared between treatment groups.

**Adverse events of study treatment:**

We hypothesize that participants randomized to convalescent plasma will not have any increased risk to standard transfusions. Preliminary data from the expanded access protocol for treatment of COVID-19 by convalescent plasma among those with severe disease, suggests that the treatment is safe with low number of adverse events [Joyner et al, J Clin Invest. 2020].

Adverse event data will be collected continuously throughout the trial and analyzed by treatment group at each interim analysis. We will also compare the rates of all serious adverse events (as specified by the Health and Human Services definition) using cumulative incidence. The primary safety endpoints are i) the cumulative incidence of treatment-related serious adverse events categorized separately as either severe infusion reactions or Acute Respiratory Distress Syndrome (ARDS) during the study period and ii) cumulative incidence of treatment-related grade 3 and 4 adverse events during the study period.

Data collection for adverse events will be done via a combination of specific questions for anticipated effects and spontaneous participant report. We will present overall number of AEs and SAEs by treatment group and where differences exist, we will present those by organ system. As these are descriptive safety analyses no correction for multiplicity will be undertaken.

**Interim monitoring:**

A multidisciplinary DSMB that will be responsible for the protection of the safety of participants enrolled in the trial. The DSMB will adopt a charter describing its responsibilities and operating characteristics. The DSMB will review the accumulating data on the primary outcome measure (SARS-CoV-2 positivity). The overall event rate (i.e., not separated by treatment arms) will be used by the DSMB to help make recommendations to mitigate risk to not successfully completing trial by providing suggestions about modifying the trial. Tracking overall event rate allows for suggestions to be made without spending alpha by conducting an interim efficacy analysis (with the exception of the planned interim analysis).

There will be two formal reviews of interim efficacy results. The first will occur after 15 COVID-19 (symptoms compatible with infection and/or RT-PCR positive) occur (after about 150 individuals) and the second after 50 percent of the randomized participants have completed 28-day follow-up data collection. Interim analysis will be adjusted for age. Stopping guidelines will be based on O-Brien Fleming boundaries; with two planned interim look (after 15 events, after 50% reach day 28), the Z-values are 3.03 (nominal p-value of 0.0011, spent alpha 0.0011), 2.38 (0.0087, 0.008), and 1.68 (0.464, 0.408) for a one-sided test with Type 1 of 0.05, respectively.

All measures will be evaluated for outliers, and distributional assumptions will be checked to ensure applicability of the statistical procedures.

The DSMB will make recommendations to the principal investigators about continuing, modifying, or stopping the trial. The DSMB will meet periodically to review interim reports and analyses derived from the accumulating data or related findings from sources external to the trial that may be needed to make recommendations to the principal investigators regarding: 1) overall efficacy and benefit/risk ratio, 2) efficacy and benefit/risk ratios within defined subsets of participants, and 3) overall and clinic-specific performance and data quality.

**Stopping guidelines:**

The trial can be terminated for any of the following reasons:

1. One treatment arm is superior to the other with substantial confidence that exceeds the planned boundaries;
2. Neither treatment arm is significantly different from the other and the possibility of achieving a difference is less than 10% with full enrollment;
3. Side effects outweigh the potential benefits of treatment in one or both treatment arms;
4. Data quality is compromised;
5. Data accrual is too slow to finish the study in a reasonable time period after considering the pandemic course and the potential for secondary outbreaks of disease;
6. External data suggests an accepted answer to the study question and investigators are no longer conducting the study under equipoise;
7. The study question is no longer relevant to the clinical community;
8. Adherence to the treatment arms is poor, leading to poor data quality;
9. There is a loss of study resources to perform the study;
10. There is evidence of fraud or misconduct in the study.

Special attention will be given to progression to ARDS and to deaths in the HCIP treatment group given the low likelihood of these outcomes under the hypothesized treatment effect.

These stopping guidelines have been adapted from *Clinical Trials: A Methodological Perspective, 2^nd^ Edition* (Piantadosi 2005).

**References:**

Benkeser, David, Iván Díaz, Alex Luedtke, Jodi Segal, Daniel Scharfstein, and Michael Rosenblum. 2020. “Improving Precision and Power in Randomized Trials for COVID‐19 Treatments Using Covariate Adjustment, for Binary, Ordinal, and Time‐to‐event Outcomes.” *Biometrics*, September. https://doi.org/10.1111/biom.13377.

Díaz, Iván, Elizabeth Colantuoni, Daniel F. Hanley, and Michael Rosenblum. 2019. “Improved Precision in the Analysis of Randomized Trials with Survival Outcomes, without Assuming Proportional Hazards.” *Lifetime Data Analysis* 25 (3): 439–68. https://doi.org/10.1007/s10985-018-9428-5.

Díaz, Iván, Elizabeth Colantuoni, and Michael Rosenblum. 2016. “Enhanced Precision in the Analysis of Randomized Trials with Ordinal Outcomes.” *Biometrics* 72 (2): 422–31. https://doi.org/10.1111/biom.12450.

**CONSORT Flow Diagram Frame**

Assessed for eligibility (n= )

#### Screening

Excluded (n=	)

-  Not meeting inclusion criteria (n= )

-  Declined to participate (n= )

-  Other reasons (n= )

#### Enrollment

Randomized (n= )

#### Allocation

Randomized to control (SARS-CoV-2 non-immune) plasma (n= )

- Received control plasma (n= )

- Received HCIP (n= )

- Did not receive transfusion (n= )

Randomized to human coronavirus immune plasma (HCIP) (n= )

- Received HCIP (n= )

- Received control plasma (n= )

- Did not receive transfusion (n= )

#### Follow-Up

#### Analysis

Analysed (n= )
- Excluded from analysis (with reasons) (n= )

Lost to follow-up (with reasons) (n= )

Analysed (n= )
- Excluded from analysis (with reasons) (n= )

Lost to follow-up (with reasons) (n= )

**Blood banking staff**

JHU

Christi Marshall, Rivcah Davis, Melissa Neally, Kristen Buban

Anne Arundel Medical Center

Sanford Robbins M.D., Megan Frisk M.S., MLS (ASCP) CM SBBCM

Baylor College of Medicine

Meredith Reyes, M.D., Laura Korte, MBA,MT(ASCP)SBB, CCRP and Benjarat Nguyen

Jackson Memorial

Yanyun Wu M.D. and Mario Charles MT

Georgetown University

James W. Reep, Albert F. Langeberg and Helen Tilahun

Lifespan/Brown University: Rhode Island Hospital

Lisa Tingley, Tracey Cheves and Karen King

NorthShore University Health System

Thomas Gniadek M.D. Ph.D. Gregory Wright MT(ASCP) SBB, CQA(ASQ)

Mayo Clinic

Jill Adamski, M.D., Ph.D.

UCLA

Alyssa Ziman M.D, Bridget Wallace MT(ASCP), SBB, Andrea McGonigle, Zhen Mei, Dawn Ward

University of Alabama at Birmingham

Marisa Marques M.D, Tammy Gray CLS , Ashton Kornbrust, Sarah Herring, Benjamin Lenard

University of California, Irvine

Minh-Ha Tran M.D, Belinda Kluchnikov MT, Marcela Feria II, Faten Mahmoud, Melissa Morris

University of California, San Diego

Angela Varela MT (ASCP), CLS, Maria (Pia) Barron, Dominga Espinoza

University of Cincinnati Medical Center

David OH, M.D. and Laura Demartino MT(ASCP) SBBcm

University of Massachusetts Worcester

Yong (David) Zhao M.D. and Natalie Malvick

University of Miami-Jackson Memorial

Yanyun Wu M.D. Alfredo Pedro Alonso, BSN, CCRC

University of New Mexico

Eileen Sierra and Cindy Jones CRN

University of Rochester

Neil Blumberg M.D, Debra Masel MT, Diane Bullock, Sarah Magri, Aimee Kievitt

University of Texas Health Science Center at Houston

Rhonda Hobbs BS MT(ASCP)SBB and Veronica Alvarado

University of Utah Health

Kelly Cali MT CLS and Yazmin Barrera

Vassar Brothers Medical Center

Daniel Cruser M.D, Susan Chumura MT(ASCP), Nicole Atkins, Charlotte Murray, Marie Bien-Aime, Tuyen Nguyen

Wayne State University

Tammon Nash, MD, Sondall Hulin MT, Jim Fiedor, Sophia Hadjiev,

Western Connecticut Health Network, Danbury Hospital

Nusrat Pathan M.D, Theresa Pearson MT , Bhumi Shah , Svetlana O'Connor

Western Connecticut Health Network, Norwalk Hospital

Bhavna Khandpur M.D, Rebecca Martinez, Jose Leonel Rodriquez, Jose Leonel Rodriquez

Milisha Patel, Norman Jones MT (ASCP)

1. *with surge of infections in December 2020 and hospitals becoming more full, these two were put into the clinical events scale as hospitalization equivalents
2. Positive molecular testing for SARS-CoV-2

   *with surge of infections in December 2020 and hospitals becoming more full, these two were put into the clinical events scale as hospitalization equivalents
